# NUTRITIONAL TRENDS AMONG CHILDREN UNDER THE AGE OF FIVE IN ZAMBIA: A LONGITUDINAL ANALYSIS USING ZAMBIA DEMOGRAPHIC HEALTH SURVEY (2001-2018)

**DOI:** 10.1101/2025.08.01.25332726

**Authors:** Namukolo Mukubesa, Leah Kamulaza, Mutale Sampa, Atupele Chisiza, Nasilele Amatende, Hope Sabao, Wilbroad Mutale

## Abstract

Childhood undernutrition, manifested as stunting, wasting, and underweight, remains a major public health challenge, particularly in low- and middle-income countries. In Zambia, the burden of undernutrition remains persistently high despite ongoing interventions. This study analyzed trends and determinants of nutritional status among children under five years using data from the Zambia Demographic and Health Survey (ZDHS) conducted in 2001, 2007, 2013–14, and 2018. The analysis assessed the prevalence of stunting, underweight, and wasting in children aged 0–59 months, using mean Z-scores and standard deviations. Logistic regression models were applied to identify key socio-demographic and health-related risk factors, with analyses performed in Python, accounting for survey design and weights.

Findings revealed a notable decline in malnutrition: stunting dropped from 45.6% in 2001 to 34.7% in 2018; underweight from 27.3% to 11.6%; while wasting remained stable at approximately 4%. Severe stunting and underweight also decreased significantly, whereas severe wasting fluctuated. Prevalence rates were higher when excluding children under six months (left-truncated data), suggesting possible protection from early infancy due to exclusive breastfeeding. Key predictors of malnutrition included low birth weight, poverty, regional disparities, and diarrheal episodes.

Despite progress, stunting remains a pressing concern. The higher rates observed in older infants point to the need for strengthened interventions targeting the postnatal period. Enhancing maternal and child health services, improving nutrition programs, and addressing poverty are critical to sustaining reductions in childhood malnutrition.

## 1. INTRODUCTION

Malnutrition in early childhood remains a major public health concern, shaping long-term health outcomes and economic productivity at both individual and national levels [3]. This issue is particularly pressing in Africa, where child undernutrition stands as one of the continent’s fundamental challenges to improved development, significantly slowing progress toward the goal of reducing malnutrition [8]. Millions of children worldwide experience the devastating effects of chronic and acute undernutrition, which not only increases the risk of mortality but also impairs cognitive development and physical growth. The three key indicators of undernutrition which are stunting, wasting, and underweight reflect different dimensions of nutritional deficits. Stunting results from prolonged nutritional deprivation, underweight signals, recent severe weight loss, and wasting encompasses aspects of both Ministry of Health (MOH) [18]. Children whose height-for-age Z-score falls below minus two standard deviations (−2 SD) from the median of the reference population are classified as stunted.

Wasting, on the other hand, is characterized by a swift loss of body weight and muscle tissue, resulting in a low weight-for-height. Like stunting, it is evaluated through a Z-score comparison, with children falling below minus two standard deviations (−2 SD) considered wasted. Underweight, a broader term, indicates a child has a low weight-for-age. Similar to the other indicators, underweight is assessed through a Z-score comparison, with children below minus two standard deviations (−2 SD) classified as underweight.

The prevalence of stunting is decreasing, while the prevalence of overweight is increasing worldwide [16]. In developing countries, approximately 32.0% of children under five years old are stunted, and 10.0% are wasted [18]. In 2014, 57.0% and 37.0%, 68.0%, and 28.0%, and 48.0% and 25.0% of all stunted, wasted, and overweight children under five lived in Asia and Africa, respectively [16]. Malnutrition is responsible for at least 35.0% of deaths among children under five worldwide [14], and nearly half of under-five deaths can be attributed to undernutrition [17].

In Zambia, despite sustained efforts to address child malnutrition, progress has been uneven. While stunting has declined moderately over the past decades, wasting and underweight prevalence rates have shown inconsistent trends, indicating persistent nutritional challenges. Similar patterns have been observed in other sub-Saharan African countries. For instance, a multi-country analysis by [1] found that while stunting has declined in some regions, wasting and underweight remain prevalent due to food insecurity, poor maternal education, and inadequate health services [1]. Existing research often focuses on cross-sectional estimates of malnutrition without addressing potential biases introduced by left truncation, a methodological issue where children younger than six months, regardless of survival status, are excluded from analysis.

This study employs data from the Zambia Demographic and Health Surveys (ZDHS) spanning 2001 to 2018 to examine long-term trends and risk factors of malnutrition. By comparing complete datasets (0–59 months) with left-truncated datasets (6–59 months), we assess how exclusion of the youngest age group influences prevalence estimates and statistical associations. Our analysis suggests that malnutrition rates appear slightly higher in left-truncated data, reinforcing the need to account for early-life vulnerabilities in nutritional assessments. Additionally, logistic regression models identify key risk factors, providing insights into socioeconomic and demographic determinants of malnutrition.

A more comprehensive understanding of malnutrition dynamics, particularly in early infancy, is crucial for designing targeted interventions. This study highlights the importance of strengthening early nutritional programs, refining assessment methodologies, and addressing gaps in policy to mitigate the long-term consequences of childhood malnutrition in Zambia.

## 2. MATERIALS AND METHODS

### 2.1 Dataset and study design

This study utilized nationally representative data on malnutrition from the ZDHS for the years 2001, 2007, 2013–14, and 2018. The ZDHS is a comprehensive household survey conducted periodically to collect data on health and demographic indicators, including child nutrition. These datasets provide critical insights into malnutrition trends among children under the age of five in Zambia.

For this analysis, we focused on the child’s recode file, which contains detailed information on child health, nutritional status, and socio-demographic characteristics. The initial sample sizes for the respective survey years were 6,031 (2001), 5,951 (2007), 12,040 (2013–14), and 9,959 (2018). After excluding cases with missing values on key anthropometric and demographic variables, the final analytical samples were reduced to 5,598 (2001), 5,351 (2007), 11,555 (2013–14), and 8,808 (2018).

The study employed a longitudinal approach to examine trends and determinants of malnutrition over time. Descriptive statistics, trend analyses, and regression models were used to assess changes in malnutrition prevalence and identify key determinants influencing child nutritional outcomes over the study period.

### 2.2 Response variable and explanatory variables

In this study, we examined three types of response variables: stunting, underweight and wasting. These variables were assessed using height-for-age Z-score (HAZ), weight-for-height Z-score (WHZ), and weight-for-age Z-score (WAZ). Z-scores were calculated utilizing World Health Organization (WHO) AnthroPlus, with age, weight, and height as determinants. Stunting was identified if HAZ fell below -2 standard deviations (SD), while wasting and underweight were characterized by WHZ and WAZ below -2 SD, respectively.

Various socio-economic and demographic factors were selected as explanatory variables, drawing from existing literature. Detailed descriptions of these variables, along with their respective categories, are presented in Tables below.

### 2.3 Statistical Analysis

#### 2.3.1 Identification of Determinant of Undernutrition

To identify key determinants of undernutrition, preliminary analyses were first conducted using chi-square tests for categorical variables and independent t-tests for continuous variables, comparing means between affected and non-affected groups. These tests helped identify statistically significant associations and informed the selection of variables for the regression model.

Following this, logistic regression analysis was employed to assess the relationship between potential risk factors and the three primary undernutrition indicators: stunting, wasting, and underweight. Logistic regression was chosen for its suitability in modeling binary outcome variables, enabling the estimation of adjusted odds ratios (AORs) with 95% confidence intervals to quantify the strength and direction of associations. All statistical analyses were performed using Python, leveraging its robust libraries for data preprocessing, modeling, and visualization.

### 2.4 Trend Analysis

To assess the trends of undernutrition among children under five in Zambia, a comprehensive trend analysis was performed using ZDHS data from 2001, 2007, 2013–14, and 2018. The study examined the prevalence of stunting, wasting, and underweight, alongside their severe forms (severe stunting, severe wasting, and severe underweight), to identify progress, emerging patterns, and persistent challenges in child malnutrition.

To provide a deeper understanding of malnutrition dynamics, the analysis was stratified into two age groups, 0–59 months which represents all children under five, offering a comprehensive view of undernutrition prevalence while 6–59 months excludes infants below six months to control for factors like exclusive breastfeeding and neonatal conditions, in order providing a clearer picture of malnutrition trends among older children.

To effectively illustrate changes over time, line graphs and bar charts were generated using Python’s Matplotlib and Seaborn libraries. Additionally, statistical trend tests were applied to determine the significance of observed patterns. These analyses provided valuable insights into long-term nutritional changes, disparities between age groups, and areas requiring targeted interventions.

#### Data Access and Ethical Considerations

The study utilized retrospective, publicly available data from the Zambia Demographic and Health Surveys (ZDHS) conducted in 2001–02, 2007, 2013–14, and 2018. Access to the datasets was granted by The DHS Program upon request through their official portal (https://dhsprogram.com). Data for this study were downloaded and accessed for analysis between **March 10 and March 15, 2024**. All datasets were fully anonymized, and the authors did not have access to any personally identifiable information at any stage of the analysis.

## 3. RESULTS

This chapter presents a comprehensive analysis of the anthropometric trends in malnutrition among children aged 0 to 59 months in Zambia. The findings focus on the global and severe prevalence of stunting, underweight, and wasting from 2001 to 2018. The chapter includes visual representations line graphs and bar charts to illustrate these trends. Further analyses explore the impact of left-truncating the dataset at 6 months (i.e., focusing on children aged 6–59 months), demographic and baseline characteristics of the sample population, and multivariate risk factors associated with malnutrition, based on logistic regression models.

### 3.1 Trends in Anthropometric Indicators of Malnutrition (0–59 Months)

The analysis of anthropometric trends among Zambian children aged 0 to 59 months from 2001 to 2018 reveals notable progress in reducing malnutrition, particularly in stunting and underweight. As shown in Table 1, stunting, measured by the HAZ, has declined from a prevalence of 45.6% in 2001 to 34.7% in 2018. Concurrently, the mean HAZ improved from -1.89 to -1.48, suggesting that, on average, children’s height-for-age has improved significantly over the 17-year period. However, despite these improvements, stunting remains a major public health concern, with over one-third of Zambian children still affected as of 2018.

**Table 1:**
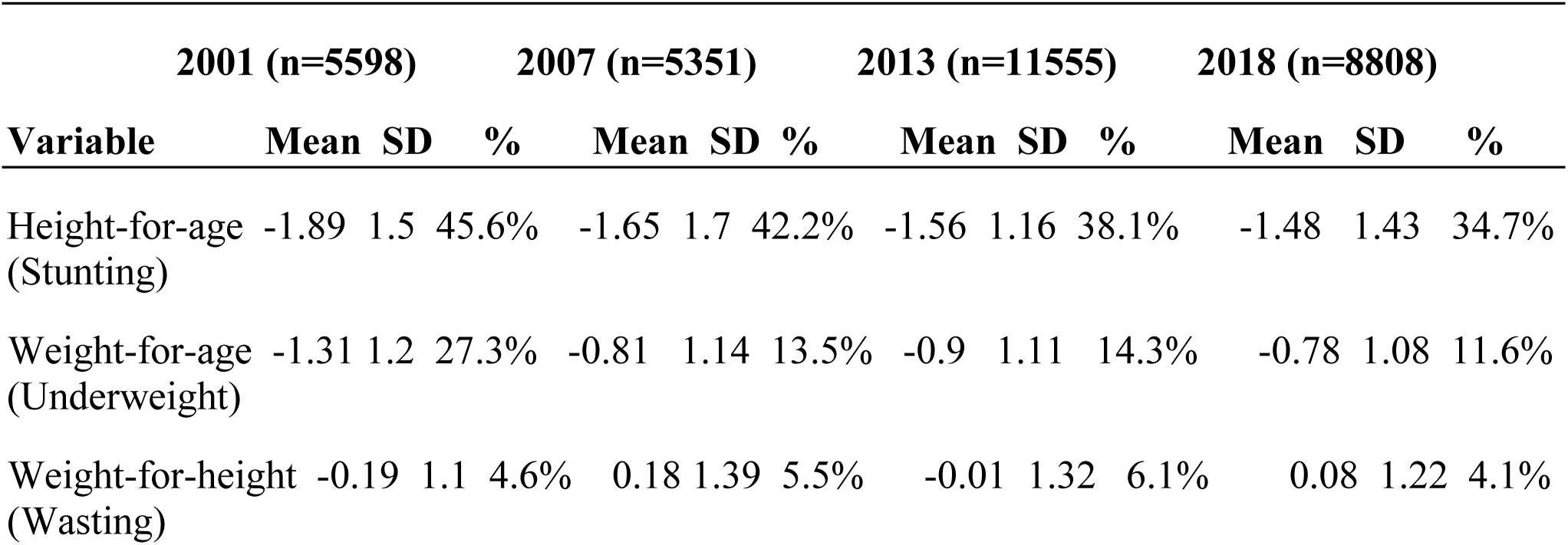
Trends of anthropometric indicators in children age 0 to 59 months (2001 to 2018) Zambia

Underweight, measured by the WAZ, also showed a substantial decline in prevalence, from 27.3% in 2001 to 11.6% in 2018. The mean WAZ increased from -1.31 to -0.78, indicating overall improvement in children’s weight-for-age. The most dramatic reduction occurred between 2001 and 2007, with the prevalence dropping from 27.3% to 13.5%, likely reflecting effective nutrition and health interventions during that period.

Wasting, measured by the WHZ, exhibited relatively stable trends with minor fluctuations. The prevalence ranged from 4.6% in 2001 to 4.1% in 2018. The mean WHZ showed variations over time, increasing to 0.18 in 2007, declining to -0.01 in 2013, and slightly recovering to 0.08 in 2018. This pattern suggests that acute malnutrition has remained relatively stable, with limited progress compared to stunting and underweight.

### 3.2 Global and Severe Prevalence of Stunting, Underweight, and Wasting

Table 2 provides insight into both global and severe forms of malnutrition from 2001 to 2018. In 2001, the global prevalence of stunting stood at 45.6%, with severe stunting affecting 21.6% of children. By 2018, stunting had declined to 34.7%, while severe stunting dropped more substantially to 11.5%. This indicates meaningful progress in reducing the extreme forms of growth retardation.

**Table 2:** Trends in Global and Severe Prevalence of stunting, underweight and wasting in children age 0 to 59 months

| Survey | Stunting |  | Underweight |  | Wasting |  |
| --- | --- | --- | --- | --- | --- | --- |
|  | Global | Severe | Global | Severe | Global | Severe |
| 2001 | 45.6% | 21.6% | 27.3% | 7.0% | 4.6% | 1.0% |
| 2007 | 42.2% | 19.5% | 13.5% | 2.8% | 5.5% | 2.1% |
| 2013-14 | 38.1 % | 16.3% | 14.3% | 3.1% | 6.1% | 2.2% |
| 2018 | 34.7% | 11.5% | 11.6% | 2.1% | 4.1% | 1.4% |

The prevalence of underweight children also improved significantly. In 2001, 27.3% of children were underweight, with 7.0% classified as severely underweight. By 2018, the overall prevalence had reduced to 11.6%, and severe underweight dropped to 2.1%, reflecting improved nutritional outcomes and likely better access to food and healthcare.

Wasting followed a less consistent pattern. The global prevalence of wasting was 4.6% in 2001 and declined slightly to 4.1% in 2018. However, severe wasting showed a modest increase from 1.0% in 2001 to 1.4% in 2018. These findings suggest that while acute malnutrition remained relatively low, progress in reducing its severe forms has been limited.

### 3.3 Visual Representation of Anthropometric Trends (2001–2018)

Figure 1 visually summarizes the overall trends in stunting, underweight, and wasting among children from 2001 to 2018. The line graph shows a steady decline in stunting, from 45.6% in 2001 to 34.7% in 2018, reflecting sustained progress in addressing chronic malnutrition. Underweight prevalence declined sharply between 2001 and 2007, from 27.3% to 13.5%, but the rate of improvement slowed thereafter, reaching 11.6% in 2018. In contrast, wasting displayed a fluctuating trend, with the highest prevalence recorded in 2013 at 6.1%, before decreasing to 4.1% in 2018. These patterns highlight the differing dynamics of chronic versus acute malnutrition, with stunting and underweight showing clearer improvement trajectories.

**Figure 1:**
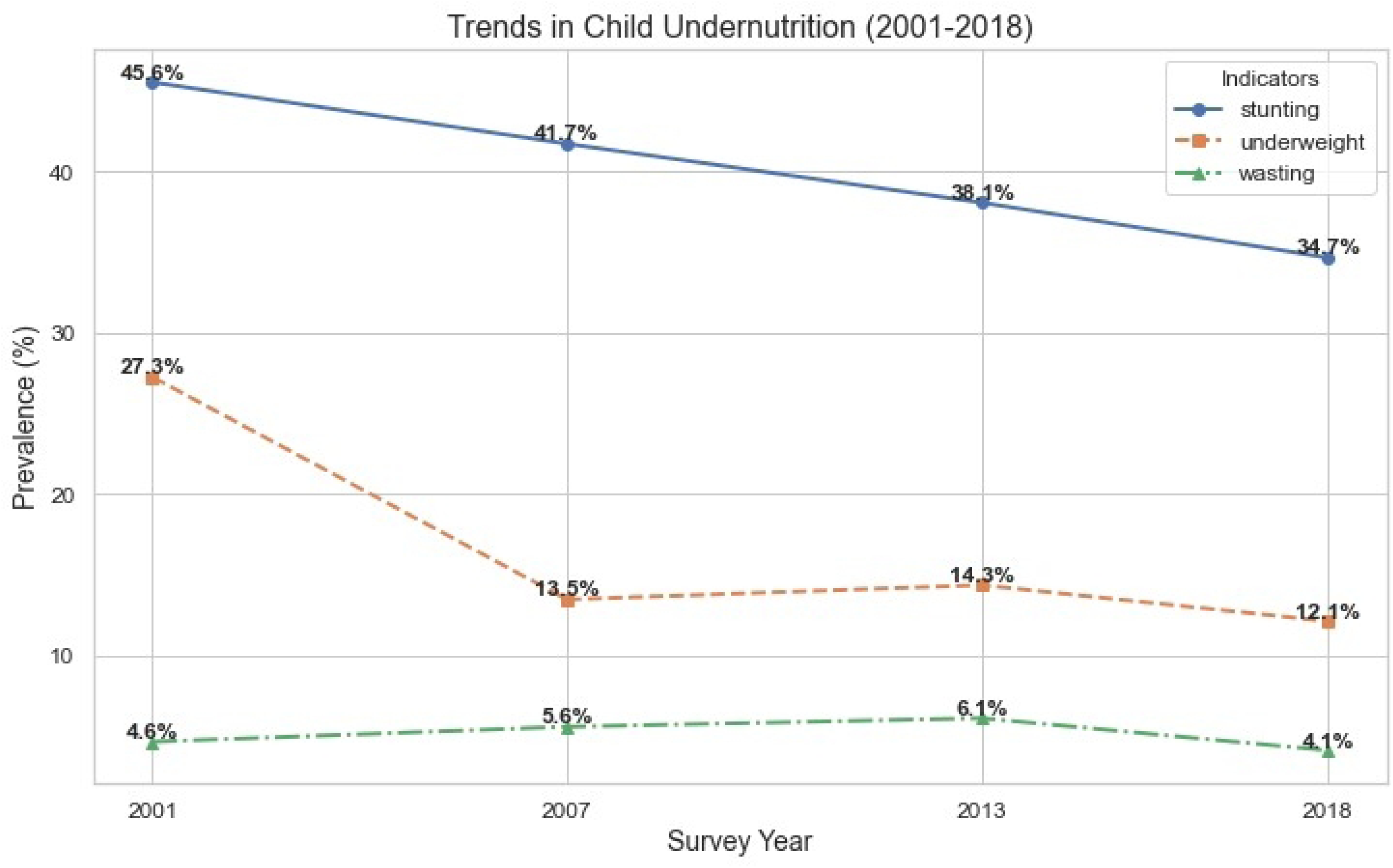
Trends in undernutrition for children under the age of 5 from 2001 to 2018

### 3.4 Left-Truncated Anthropometric Indicators (6–59 Months)

Table 3 presents anthropometric trends when data are restricted to children aged 6 to 59 months, allowing for comparison with the full 0–59 months sample. The results from Figures 2, 3, and 4 reveal that stunting and underweight prevalence were consistently higher among children aged 6– 59 months than those aged 0–59 months. In 2001, for example, stunting increased from 45.6% to 50.4%, and underweight rose from 27.3% to 30.4% when children under six months were excluded. This indicates that younger infants typically exhibit lower levels of chronic malnutrition.

**Table 3:** Prevalence of stunting, underweight and wasting when the data was left-truncated at 6 months (6-59).

| Survey | Stunting |  | Underweight |  | Wasting |  |
| --- | --- | --- | --- | --- | --- | --- |
|  | Global | Severe | Global | Severe | Global | Severe |
| 2001 | 50.4% | 24.2% | 30.4% | 7.9% | 4.9% | 1.0% |
| 2007 | 44.1% | 20.6% | 14.1% | 3.0% | 5.4% | 2.1% |
| 2013-14 | 40.7 % | 17.6% | 15.3% | 3.3% | 5.6% | 2.1% |
| 2018 | 36.5% | 12.3% | 12.5% | 2.4% | 4.0% | 1.3% |

**Figure 2:**
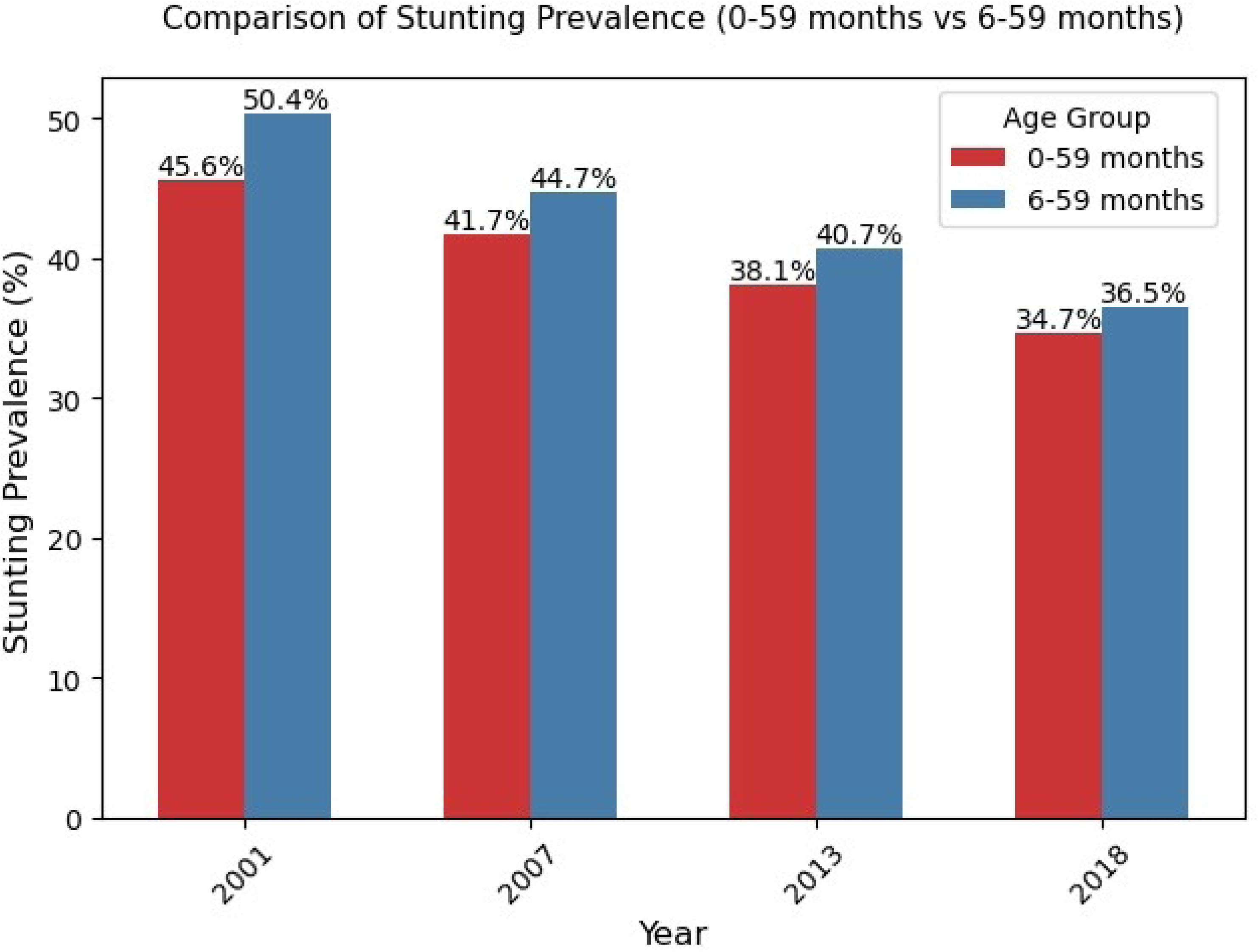
Comparison of stunting prevalence between complete dataset ( 0 -59 months) verses left-truncated dataset (6 – 59 months)

**Figure 3:**
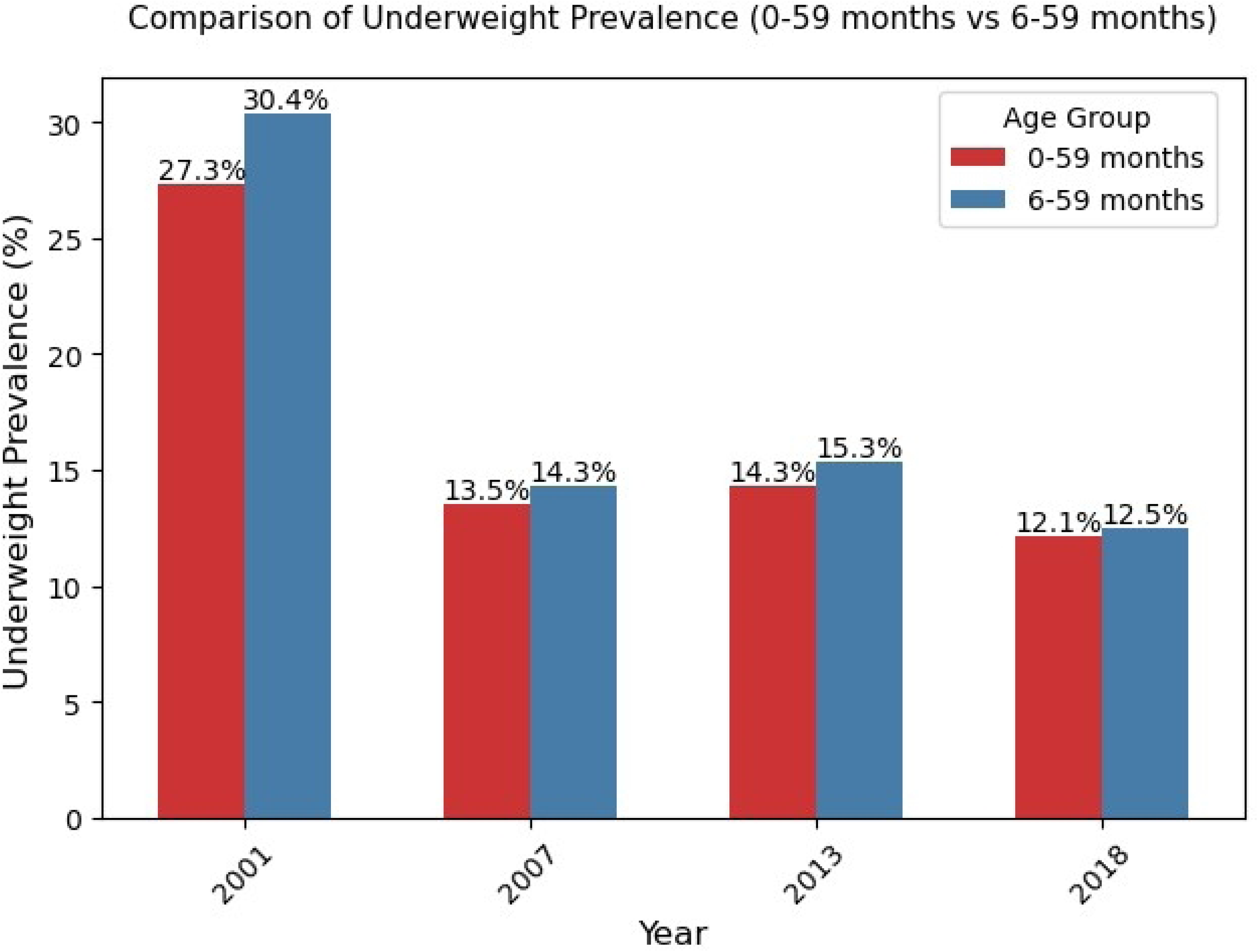
Comparison of underweight prevalence between complete dataset ( 0 -59 months) verses left-truncated dataset (6 – 59 months)

**Figure 4:**
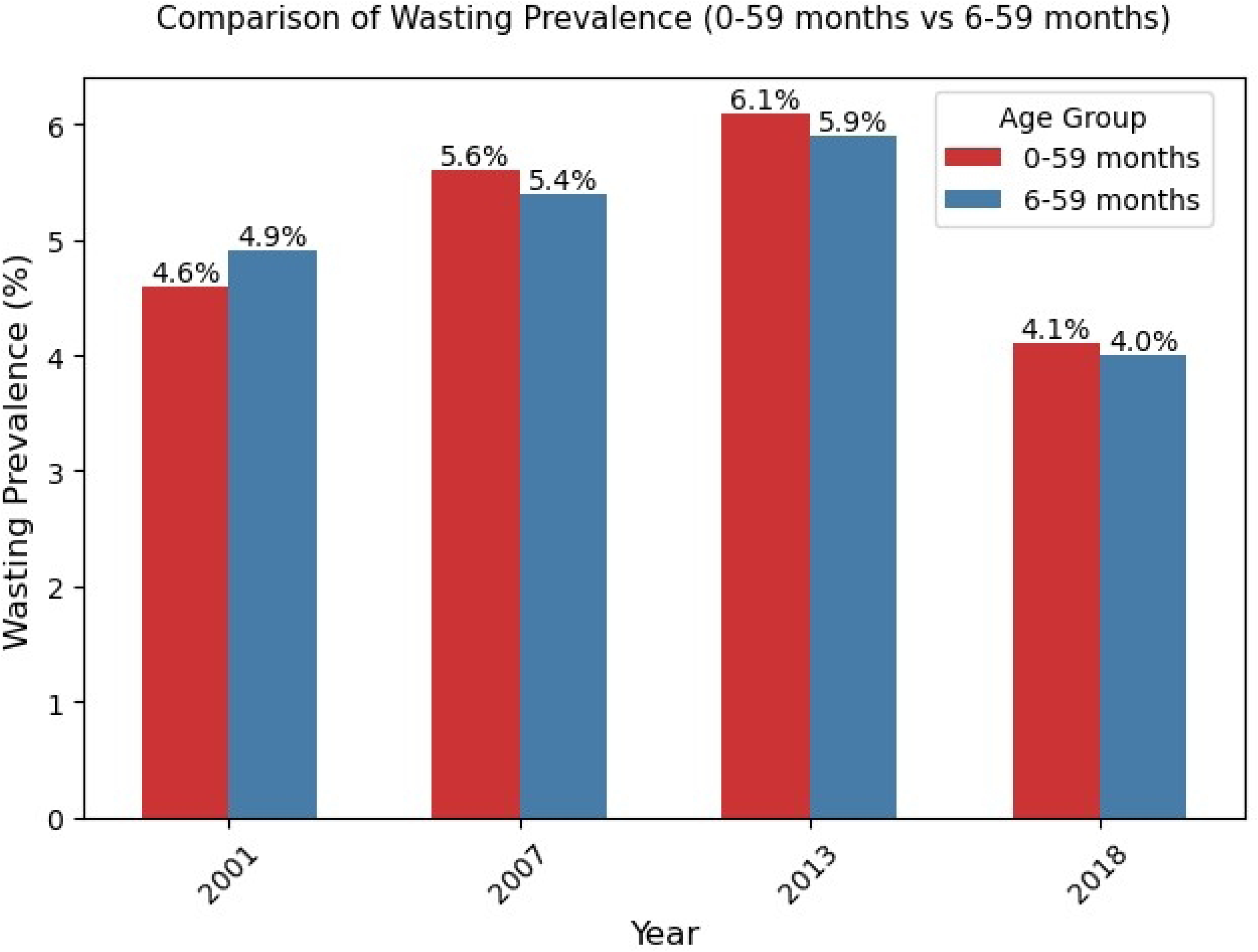
Comparison of wasting prevalence between complete dataset ( 0 -59 months) verses left-truncated dataset (6 – 59 months)

Interestingly, the wasting indicator behaved differently. The exclusion of children aged 0–5 months had only a marginal effect on the prevalence of wasting. For instance, in 2007, wasting declined slightly from 5.5% to 5.4% when the data were left-truncated at six months. This minimal change suggests that wasting in infants may be influenced by different factors than stunting and underweight, warranting further investigation.

### 3.5 Baseline and Demographic Characteristics of Respondents

Table 4 presents a summary of the baseline and demographic characteristics of survey respondents, using combined data from the 2001, 2007, 2013–2014, and 2018 ZDHS. Due to variable unavailability, the wealth index data were derived only from 2007, 2013, and 2018 datasets, and provincial data were combined from 2013 and 2018, as Muchinga Province was not included in earlier surveys.

**Table 4:** Baseline and demographic characteristic of respondents for stunting, wasting and underweight variables

| Variables | Stunted | P-value | Wasting | P-value | Underweight | P-value |
| --- | --- | --- | --- | --- | --- | --- |
| Respondent age group |  | <0.05 |  | 0.43 |  | 0.3 |
| 25-34 | 38.5% (5513) |  | 5% (717) |  | 15.7% (2244) |  |
| <=24 | 40.5% (4129) |  | 5.2% (533) |  | 16.3% (1665) |  |
| >=35 | 38.1% (2564) |  | 5.4% (364) |  | 15.6% (1050) |  |
| Region |  | <0.01 |  | <0.01 |  | <0.01 |
| Central | 36.6% (667) |  | 4.3% (79) |  | 12.5% (227) |  |
| C/belt | 32.9% (600) |  | 5% (92) |  | 13.3% (243) |  |
| Eastern | 37% (901) |  | 4.9% (119) |  | 10.1% (246) |  |
| Luapula | 41.8% (970) |  | 8.8% (203) |  | 17.4% (404) |  |
| Lusaka | 33.5% (656) |  | 6.3% (123) |  | 10.6% (207) |  |
| Muchinga | 37.8% (731) |  | 5.7% (110) |  | 15.1% (291) |  |
| N/Western | 33.9% (653) |  | 5.3% (103) |  | 12.5% (240) |  |
| Northern | 45.6% (1033) |  | 3% (69) |  | 16.2% (368) |  |
| Southern | 32% (693) |  | 3.5% (76) |  | 11.1% (240) |  |
| Western | 31.6% (522) |  | 5.1% (84) |  | 15% (247) |  |
| Size of child |  | <0.01 |  | <0.05 |  | <0.01 |
| Average | 39.3% (7358) |  | 5% (938) |  | 16% (2995) |  |
| large | 34.2% (2944) |  | 4.8% (416) |  | 11.3% (973) |  |
| small | 48.7% (1904) |  | 6.7% (260) |  | 25.4% (991) |  |
| Wealth Index |  | <0.01 |  | <0.05 |  | <0.01 |
| Middle | 37.5% (2092) |  | 4.5% (252) |  | 12.2% (683) |  |
| Poor | 41.9% (5256) |  | 5.6% (698) |  | 15.8% (1985) |  |
| Rich | 30.7% (2310) |  | 5.4% (405) |  | 10.2% (765) |  |
| Rural/Urban |  | <0.01 |  | 0.38 |  | <0.05 |
| Rural | 41.4% (8842) |  | 5.1% (1087) |  | 17% (3628) |  |
| Urban | 34% (3364) |  | 5.3% (527) |  | 13.5% (1331) |  |
| Education Level |  | <0.01 |  | 0.8 |  | <0.01 |
| Higher | 16.6% (156) |  | 4.9% (46) |  | 6.8% (64) |  |
| No-edu | 42.8% (1623) |  | 6% (229) |  | 20.3% (770) |  |
| Primary | 42% (7574) |  | 5% (898) |  | 17% (3062) |  |
| Secondary | 33.7% (2853) |  | 5.2 (441) |  | 12.66 (1063) |  |
| Mother work status |  | <0.01 |  | 0.65 |  | <0.05 |
| No | 37.8% (5389) |  | 5.1% (727) |  | 15.3% (2179) |  |
| Yes | 40.1% (6817) |  | 5.2% (887) |  | 16.4% (2780) |  |
| Sex of child |  | <0.01 |  | <0.05 |  | <0.05 |
| F | 36.7% (5746) |  | 4.7% (736) |  | 14.6% (2292) |  |
| M | 41.4% (6460) |  | 5.6% (878) |  | 17.1% (2667) |  |
| Toilet facility |  | <0.01 |  | 0.39 |  | <0.05 |
| Other | 43.2% (4049) |  | 4.9% (460) |  | 20.9% (1959) |  |
| Pitlatrine | 38.4% (6719) |  | 5.3% (922) |  | 13.6% (2380) |  |
| Watercloset | 32.9% (1438) |  | 5.3% (232) |  | 14.2% (620) |  |
| Age first preg in yrs | 18 (3.04) | <0.01 |  | 0.08 |  | <0.05 |
| Breastfeeding status |  | <0.01 |  | <0.05 |  | 0.135 |
| No | 41% (5388) |  | 4.5% (595) |  | 15.5% (2037) |  |
| Yes | 37.7% (6818) |  | 5.6% (1019) |  | 16.1% (2922) |  |
| Water source |  | <0.01 |  | 0.69 |  | <0.05 |
| Borehole | 36.7% (1817) |  | 4.9% (244) |  | 13.1% (651) |  |
| Other | 37.1% (280) |  | 5.4% (41) |  | 15.8% (119) |  |
| Tap | 33.1% (2489) |  | 5.4% (404) |  | 13.4% (1004) |  |
| Well | 42.3% (7620) |  | 5.1% (925) |  | 17.7% (3185) |  |
| Diarrhea |  | <0.01 |  | <0.05 |  | <0.05 |
| No | 38.3% (9918) |  | 4.9% (1256) |  | 14.8% (3823) |  |
| Yes | 42.5% (2288) |  | 6.7% (358) |  | 21.1% (1136) |  |
| Fever |  | <0.01 |  | 0.18 |  | <0.05 |
| No | 38.1% (9035) |  | 5.1% (1201) |  | 14.2% (3372) |  |
| Yes | 41.9% (3171) |  | 5.5% (413) |  | 21% (1587) |  |
| Cough |  | 0.76 |  | 0.44 |  | <0.05 |
| No | 39% (9019) |  | 5.2% (1208) |  | 15.2% (3506) |  |
| Yes | 39.1% (3187) |  | 5% (406) |  | 17.9% (1453) |  |

Regionally, Northern province exhibited the highest stunting prevalence (45.6%), while Western province recorded the lowest (31.6%), indicating regional disparities in nutritional outcomes. Luapula province had the highest prevalence of wasting at 8.8%, pointing to a higher burden of acute malnutrition, whereas Northern province had the lowest wasting prevalence (3.0%). Underweight prevalence was highest in Luapula (17.4%) and lowest in Eastern province (10.1%). These differences highlight the need for region-specific interventions.

Birth size was strongly associated with malnutrition. Children born small or very small were more likely to be stunted (48.7%), underweight (25.4%), or wasted (6.7%) than those born average or larger in size. Rural residence was also linked to worse outcomes, with stunting rates at 41.4% compared to 34.0% in urban areas. Maternal education was inversely related to stunting; mothers with no education had a stunting prevalence of 42.8%, while those with higher education had only 16.6%. Boys were more affected by stunting (41.4%) than girls (36.7%), suggesting possible gender-related vulnerability.

Several other factors were associated with undernutrition. These include region, residence, mother’s education, household wealth, toilet and water access, breastfeeding practices, and recent illness episodes such as diarrhea. Maternal age at first pregnancy and the sex of the child were also significant predictors of undernutrition. The multifactorial nature of these associations underscores the importance of holistic, integrated approaches to combat malnutrition.

### 3.6 Logistic Regression Analysis of Risk Factors

Table 5 presents the results of a logistic regression analysis on the risk factors for stunting, underweight, and wasting. Significant regional differences are observed. The odds of stunting are 14%, 17%, and 32% lower for children in Northwestern, Southern, and Western provinces, respectively, compared to Central province. Conversely, the odds of underweight are 14% and 28% higher in Luapula and Northern provinces, respectively. For underweight, children in Eastern province have a 28% lower risk, while those in Copperbelt and Luapula provinces have 24% and 36% higher odds, respectively. With reference to wasting, children in Northern province have a 35% lower risk, whereas those in Luapula and Lusaka provinces are twice as likely and 40% more likely to be wasted, respectively, compared to Central province.

**Table 5:** Risk Factors Extraction for Stunting, Underweight and Wasting Using Logistic Regression

| Variables | Stunted<br>OR (95% CI) | p-value | Underweight<br>OR (95% CI) | P-value | Wasting<br>OR (95% CI) | P-val<br>ue |
| --- | --- | --- | --- | --- | --- | --- |
| Region |  |  |  |  |  |  |
| Central | Ref |  | Ref |  | Ref |  |
| C/belt | 0.95 (0.83, 1.10) |  | 1.24 (1.10,1.52)* |  | 1.16(0.84, 1.58) |  |
| Eastern | 0.98 (0.86,1.12) |  | 0.72 (0.59, 0.88)* |  | 1.15 (0.85, 1.55) |  |
| Luapula | 1.14(1.01,1.31)* |  | 1.36 (1.13, 1.64)* |  | 2.1 (1.56, 2.72)* |  |
| Lusaka | 1.13 (0.98, 1.31) |  | 1.06 (0.86, 1.31) |  | 1.4 (1.02, 1.88)* |  |
| Muchinga | 0.98 (0.85, 1.13) |  | 1.14 (0.95, 1.39) |  | 1.28 (0.95, 1.73) |  |
| N/Western | 0.86(0.75, 0.99)* |  | 0.96 (0.79, 1.18) |  | 1.21 (0.89, 1.64) |  |
| Northern | 1.28(1.12, 1.46)* |  | 1.01 (0.97, 1.42) |  | 0.65(0.46,0.91)* |  |
| Southern | 0.83(0.73, 0.96)* |  | 0.9 (0.74, 1.10) |  | 0.81 (0.59, 1.13) |  |
| Western | 0.68(1.12, 1.46)* |  | 1.01 (0.81, 1.23) |  | 1.16 (0.83, 1.61) |  |
| Size of child |  | <0.01 |  | <0.01 |  | <0.01 |
| Average | Ref |  | Ref |  | Ref |  |
| large | 0.79 (0.75, 0.84)* |  | 0.66 (0.61, 0.72)* |  | 0.94 (0.84, 1.06) |  |
| small | 1.45 (1.35, 1.55)* |  | 1.75 (1.61,1.91)* |  | 1.35(1.17, 1.56)* |  |
| Wealth Index |  | <0.01 |  | 0.2 |  | <0.05 |
| Middle | Ref |  | Ref |  | Ref |  |
| Poor<br>Rich | 1.3 (1.15, 1.48)*<br>0.93 (0.79, 1.09) |  | 1.34 (1.07, 1.55)*<br>0.87 (0.76, 0.97)* |  | 1.25 (1.07, 1.47)<br>0.97(0.92, 1.39)* |  |
| Rural/Urban |  | <0.01 |  | <0.05 |  | 0.4 |
| Rural<br>Urban | Ref<br>0.96 (0.9, 1.03) |  | Ref<br>1.09 (1.01, 1.19)* |  | Ref<br>1.01 (0.88,1.16) |  |
| Education<br>Level |  | <0.05 |  | <0.05 |  | 0.87 |
| Higher<br>No-edu<br>Primary<br>Secondary | Ref<br>2.83(2.32, 3.44)*<br>2.81(2.33, 3.39)*<br>2.18(1.81, 2.62)* |  | Ref<br>2.43 (1.83, 3.23)*<br>2.07 (1.57, 2.72)*<br>1.63 (1.24, 2.14)* |  | Ref<br>1.35 (0.94, 1.93)<br>1.09 (0.78, 1.53)<br>1.11 (0.8, 1.55) |  |
| Sex of Child |  | <0.01 |  | <0.05 |  | <0.05 |
| F<br>M | Ref<br>1.26 (1.2, 1.32)* |  | Ref<br>1.25 (1.18, 1.34)* |  | Ref<br>1.22 (1.1, 1.35)* |  |
| BreastFeedg |  | <0.01 |  | 0.9 |  | <0.01 |
| No<br>Yes | Ref<br>0.78 (0.75, 0.82)* |  | Ref<br>0.93 (0.87, 0.99)* |  | Ref<br>1.26 (0.98, 1.4) |  |
| Mother<br>working sta<br>tus |  | <0.05 |  | <0.05 |  | 0.2 |
| No<br>Yes | Ref<br>1.07(1.02, 1.13)* |  | Ref<br>1.02 (0.96, 1.08) |  | Ref<br>1.04 (0.94, 1.15) |  |
| Toilet<br>facility |  | <0.05 |  | <0.05 |  | <0.05 |
| Other<br>Pitlatrine<br>Watercloset | Ref<br>0.88 (0.84,0.93)*<br>0.84 (0.77, 0.92)* |  | Ref<br>0.68 (0.64, 0.73)*<br>0.71 (0.62, 0.8)* |  | Ref<br>1.11 (0.98, 1.25)<br>1.1 (0.92,1.34) |  |
| Water<br>Source |  | <0.01 |  | <0.05 |  | 0.9 |
| Borehole<br>Other<br>Tap<br>Well | Ref<br>0.89 (0.75, 1.05)<br>0.79 (0.73, 0.86)*<br>1.02 (0.97, 1.08) |  | Ref<br>0.75 (0.91, 1.82)<br>0.66 (0.58, 0.76)*<br>0.84 (0.78, 1.9) |  | Ref<br>1.03 (0.72, 1.48)<br>1.16 (0.97, 1.39)<br>1.04 (0.91, 1.17) |  |
| Anemic<br>levels |  | <0.01 |  | 0.3 |  | 0.9 |
| Mild<br>Moderate<br>Not Anemic<br>Severe | Ref<br>1.15 (1.8, 1.36)*<br>0.77(0.68, 0.92)* |  | Ref<br>1.27 (1.5, 1.65)*<br>0.91 (0.79, 1.09) |  | Ref<br>0.89 (0.69, 1.24)<br>1.09 (0.79, 1.35)<br>0.89 (0.43, 2.09) |  |
|  | 1.59(1.15, 2.27)* |  | 2.12 (1.39 , 3.89)* |  |  |  |
| Age first preg in yrs | 0.98(0.95, 0.99)* | <0.01 | 0.99 (0.98, 1.01) | 0.5 | 1.01 (0.98, 1.02) | 0.5 |
| Diarrhea |  | <0.01 |  | <0.05 |  | <0.05 |
| No<br>Yes | Ref<br>1.17 (1.1, 1.24)* |  | Ref<br>1.37 (1.27, 1.49)* |  | Ref<br>1.37(1.21, 1.55)* |  |
| Fever |  | <0.01 |  | 0.21 |  | 0.5 |
| No<br>Yes | Ref<br>1.08 (1.02, 1.15)* |  | Ref<br>1.34 (1.24, 1.45)* |  | Ref<br>1.07 (0.94, 1.22) |  |
| Cough |  | 0.2 |  | 0.4 |  | 0.2 |
| No<br>Yes | Ref<br>0.93 (0.88, 0.98)* |  | Ref<br>0.97 (0.9, 1.05) |  | Ref<br>0.88 (0.78,1.01) |  |

Birth size has a significant association with all three forms of undernutrition. Children born small or very small have 45% higher odds of stunting and 75% higher odds of underweight compared to those born average or large. Wealth status also plays a role, as children from poor households have a 30% higher likelihood of being stunted than those from middle-income households.

Maternal education was a strong predictor of nutritional status. Children whose mothers had no formal education were 2.83 times more likely to be stunted and 2.43 times more likely to be underweight than those with mothers who had higher education. Male children were disproportionately affected, with boys having 26% higher odds of stunting, 25% higher odds of underweight, and 22% higher odds of wasting compared to girls.

Breastfeeding offered a protective effect against stunting, reducing its odds by 22%, although it did not significantly affect wasting. Children of working mothers had a 7% higher risk of stunting. Improved sanitation and water sources were associated with reduced undernutrition, suggesting that environmental health plays a key role.

Recent illnesses, particularly diarrhea and fever, were significantly linked to malnutrition. Children who had diarrhea in the past two weeks were 17% more likely to be stunted and 37% more likely to be underweight. Fever increased the odds of stunting by 8% and underweight by 34%, although it did not significantly influence wasting. These findings highlight the importance of infection control in reducing childhood malnutrition.

## 4. DISCUSSION

The findings of this study provide valuable insights into the trends and determinants of undernutrition among children under five in Zambia between 2001 and 2018. The results indicate significant progress in reducing stunting and underweight, while wasting has remained relatively unchanged. This pattern suggests that while long-term interventions may be mitigating chronic malnutrition, acute malnutrition persists—reflecting gaps in short-term food security and child healthcare. Similar trends have been observed in other low- and middle-income countries, such as Nigeria and Ethiopia, where stunting has declined but wasting remains prevalent due to seasonal food shortages and disease outbreaks [1] [16].

The decline in stunting from 45.6% in 2001 to 34.7% in 2018 reflects improvements in child nutrition, healthcare access, and socio-economic conditions. However, stunting remains a significant concern, still affecting more than one-third of Zambian children. These findings are consistent with national-level studies, such as that by [10], which highlighted the role of food insecurity, poor maternal health, and inadequate feeding practices. Internationally, similar associations have been documented in South Asia and sub-Saharan Africa, where maternal undernutrition and low dietary diversity are key drivers of child stunting [3].

The prevalence of underweight declined markedly from 27.3% in 2001 to 11.6% in 2018, with the most rapid improvements observed between 2001 and 2007. This aligns with Lusaka-based research associating improvements in maternal nutritional knowledge and feeding practices with reduced child underweight [11]. However, the slower decline after 2007 suggests diminishing returns from early interventions, highlighting the need for sustained investment in child nutrition programs and maternal support systems. A similar plateauing effect was observed in Kenya and Uganda, where initial nutrition programs achieved early gains that were not maintained due to funding and implementation gaps.

In contrast, wasting prevalence has fluctuated slightly, from 4.6% in 2001 to 4.1% in 2018, while severe wasting increased from 1.0% to 1.4%. This pattern reflects the episodic nature of acute malnutrition, influenced by short-term factors like disease outbreaks, seasonal food insecurity, and natural disasters. Similar findings have been reported in West African countries, where wasting rates remain stubbornly high due to climate shocks and limited access to emergency nutrition services [1].

This study also reveals significant regional disparities in nutritional outcomes. Northern Province recorded the highest stunting rate (45.6%), while Western Province had the lowest (31.6%). Luapula Province had the highest rates of both wasting (8.8%) and underweight (17.4%). These regional differences reflect disparities in food access, healthcare coverage, and household income levels, consistent with findings from Malawi and Ethiopia where rural, impoverished regions face higher undernutrition burdens [16].

Maternal education emerged as a strong protective factor. Children whose mothers had no formal education were 2.83 times more likely to be stunted than those whose mothers attained higher education. This mirrors findings from studies in Nigeria and Bangladesh, which confirm that maternal education significantly reduces the risk of child undernutrition through improved health-seeking behavior and better child feeding practices [1]. Lusaka-based research further supports this, showing that poor maternal understanding of malnutrition prevention contributes to elevated child undernutrition rates [11].

The logistic regression analysis confirms that multiple socio-economic and environmental factors significantly contribute to undernutrition. Birth size was a strong predictor: children born small or very small had higher odds of being stunted or underweight. This is consistent with findings from India and Nepal, where low birth weight is a major risk factor for poor nutritional outcomes [3]. These findings underscore the importance of maternal nutrition and antenatal care in reducing neonatal and infant health risks.

Household wealth also played a critical role. Children from poor households were 30% more likely to be stunted than those from middle-income households. This wealth disparity reflects structural socio-economic inequalities, such as food insecurity and limited access to health and social services. Similar findings have been reported in multi-country analyses across sub-Saharan Africa and South Asia [1]. Targeted poverty-reduction strategies such as cash transfers and food supplementation are essential to address these disparities.

Access to water, sanitation, and hygiene (WASH) was another important determinant. Children in households with access to improved toilet facilities and clean water were less likely to be malnourished. Poor sanitation increases exposure to infections like diarrhea, which in turn compromise nutrient absorption and immune function. These results are supported by regional studies from Ethiopia and Ghana, which found that improved WASH conditions significantly reduced child undernutrition.

Other consistent risk factors include frequent childhood illnesses, such as diarrhea and fever, which were associated with higher rates of stunting and underweight. Repeated infections increase metabolic demands and reduce nutrient absorption, exacerbating malnutrition. Similar associations have been reported in both local [11] and international studies [3]. Strengthening healthcare systems, expanding immunization coverage, and improving caregiver education on disease prevention are crucial to breaking the infection-malnutrition cycle.

## Strengths and Limitations

This study has several notable strengths. First, it uses nationally representative ZDHS data collected over nearly two decades, enabling a robust assessment of temporal trends in undernutrition among children under five in Zambia. The application of standardized anthropometric indicators and multivariate logistic regression strengthens the internal validity of the findings and supports comparisons across different regions and time points.

Second, the study uniquely compares undernutrition prevalence based on both complete data (0– 59 months) and left-truncated data (6–59 months). This distinction is important because it highlights how exclusion of younger infants who tend to have lower prevalence of stunting and underweight can inflate overall estimates. Such methodological transparency improves the interpretability of findings and has implications for survey design and policy interpretation.

Third, the analysis includes both overall and **severe forms** of stunting, wasting, and underweight, which allows for a more nuanced understanding of the severity of child undernutrition. This approach enables better alignment with global targets (e.g., WHO thresholds) and enhances the utility of the results for international benchmarking and prioritization of high-risk cases.

However, the study is not without limitations. Although the original aim was to analyze nutritional trends from 1992 to 2024, this was not feasible due to incomplete or unavailable data for the 1992, 1997, and 2024 surveys. Consequently, the analysis was limited to data from 2001 to 2018, potentially missing earlier and more recent developments.

Additionally, the cross-sectional nature of the ZDHS limits the ability to draw causal inferences. Several potentially important variables such as dietary diversity, household food insecurity, and maternal mental health were either inconsistently available or omitted from the surveys, reducing the ability to assess their influence. Further, self-reported measures such as child illness and birth size may be subject to recall and reporting bias. Lastly, unobserved confounders, such as intra-household food allocation and caregiving practices, may also affect child nutrition outcomes but were not captured in the dataset.

## Policy Implications

The findings have several policy implications for Zambia’s ongoing efforts to reduce child undernutrition:

- **Strengthen maternal education programs** by integrating nutrition education and health literacy into both formal and informal settings.
- **Expand social protection measures,** such as targeted cash transfers and food support, especially for rural and low-income households.
- **Enhance WASH infrastructure** to reduce infection-related malnutrition, particularly in underserved regions.
- **Invest in maternal and child healthcare**, including prenatal care and community-based growth monitoring, to prevent low birth weight and promote early detection of undernutrition.
- **Ensure sustainable funding** for national nutrition programs to avoid stagnation in progress, especially for reducing wasting.

## Future Research Directions

Future studies should explore the following areas:

- **Longitudinal studies** to assess causal pathways and the long-term impact of early-life undernutrition.
- **Qualitative research** to understand community-level barriers to maternal and child nutrition.
- **Evaluation of specific interventions,** such as community-based nutrition programs or emergency feeding schemes, to identify the most effective strategies.
- **Analysis of emerging trends,** such as urban undernutrition or the impact of climate change on child health, using more recent ZDHS or other national survey data.

## 5. CONCLUSION

Although Zambia has made commendable progress in reducing stunting and underweight, undernutrition remains a pressing public health concern. The persistent regional and socio-economic disparities highlight the need for targeted interventions that address localized challenges. Comparisons with previous studies reinforce that multiple factors including maternal education, household wealth, healthcare access, and sanitation play significant roles in child nutritional outcomes. A comprehensive, multi-sectoral approach that integrates nutrition, healthcare, education, and social protection is essential to achieving sustainable improvements in child nutrition and ensuring that all Zambian children have a healthy start in life.

### What Is Already Known On This Topic

Several studies have analyzed the prevalence of stunting, wasting, and underweight at each data points, consistently highlighting that malnutrition among children under five remains a major public health concern.

It is also well established in the literature that the burden of malnutrition is not evenly distributed across countries or regions. In Zambia, national surveys like the ZDHS have consistently shown high levels of stunting, particularly in rural provinces such as Northern and Luapula, pointing to persistent regional inequalities in nutritional outcomes. For instance, [3] and UNICEF reports have emphasized that socioeconomic factors, maternal education, and access to health services contribute significantly to these disparities.

Additionally, most previous studies on child nutrition in Zambia and similar settings have used the full sample of children aged 0–59 months when reporting prevalence rates. Few have explored how prevalence estimates might shift if different age sub-groups such as excluding children under six months.

### What This Study Adds

This study adds new insights into how the choice of age group influences the measurement and interpretation of child malnutrition in Zambia. By comparing the full dataset (children aged 0–59 months) with a truncated dataset (children aged 6–59 months), we reveal that excluding the youngest infants who are often exclusively breastfed and less exposed to inadequate complementary feeding leads to higher prevalence estimates for both global and severe forms of stunting and underweight. This pattern was consistent across all four DHS survey years (2001/2, 2007, 2013/14, and 2018), highlighting that age-based inclusion criteria can significantly affect the perceived burden of malnutrition.

In contrast, wasting showed a slight decrease in the truncated sample, while severe wasting increased marginally, illustrating the different age-related patterns of acute versus chronic malnutrition. By disaggregating both global and severe forms of each indicator, our study provides a more nuanced understanding of nutritional trends, showing that while overall levels of severe stunting dropped from 21.6% to 11.5% over the study period, severe forms still represent a substantial public health concern.

Additionally, our study confirm that boys are consistently more affected than girls across all three indicators offering robust local evidence that supports global patterns and strengthens the case for gender-sensitive nutrition interventions in Zambia. Overall, this study emphasizes that methodological decisions especially regarding age stratification and the inclusion of severity levels can shape not only prevalence estimates but also policy priorities.

Our study informs policymakers by providing a more detailed understanding of childhood malnutrition in Zambia through regional and trend analysis, as well as by highlighting the importance of disaggregating severity levels in stunting, wasting, and underweight. By demonstrating that excluding children under six months yields higher prevalence estimates particularly for stunting and underweight we show how methodological decisions can significantly affect the interpretation of nutritional data. The consistent finding that boys are more affected than girls also strengthens the case for gender-sensitive nutrition interventions. These insights support the need for more targeted, age-specific, and regionally responsive policies to effectively address both chronic and acute forms of child undernutrition.

## Authors’ Contributions

Namukolo Mukubesa led the conceptualization and design of the study, collected the datasets, conducted all data analyses, and drafted the initial manuscript. Leah Kamulaza contributed to the study design and provided overall supervision of the project. All authors critically reviewed the manuscript for intellectual content and approved the final version for submission.

## Data Availability

All data underlying the findings described in this manuscript are fully available without restriction from the Demographic and Health Surveys (DHS) Program. The datasets used in this study (ZDHS 2001-2002, 2007, 2013-14, and 2018) are accessible by registered users at the DHS Program website: https://dhsprogram.com/data/available-datasets.cfm. Researchers can request access by registering and stating the purpose of data use.

## Acknowledgements

We would like to express our deepest gratitude to all those who supported us throughout this research journey. We are also grateful to the Zambia Statistics Agency (ZamStats) and the Demographic and Health Surveys Program for granting access to the Zambia Demographic and Health Survey (ZDHS) datasets used in this study.

## Tables and Figures

## REFERENCES

1. Akombi B, Agho K, Merom D, Hall J, Renzaho A. Multilevel analysis of factors associated with wasting and underweight among children under-five years in Nigeria. Nutrients. 2017;9(1):44. **<u>PubMed</u> | <u>Google Scholar</u>**

2. Black RE, Allen LH, Bhutta ZA, et al. Maternal and child undernutrition: global and regional exposures and health consequences. Lancet. 2008;371:243–60. **<u>PubMed</u> | <u>Google</u> <u>Scholar</u>**

3. Black RE, Victora CG, Walker SP, et al. Maternal and child undernutrition and overweight in low-income and middle-income countries. Lancet. 2013;382:427–51. **<u>PubMed</u> | <u>Google</u> <u>Scholar</u>**

4. Bloss E, Wainaina F, Bailey RC. Prevalence and predictors of underweight, stunting and wasting among children aged 5 and under in Western Kenya. J Trop Pediatr. 2004;50 (5):260–70. **<u>PubMed</u> | <u>Google Scholar</u>**

5. Caulfield LE, de Onis M, Blössner M, Black RE. Undernutrition as an underlying cause of child deaths associated with diarrhea, pneumonia, malaria and measles. Am J Clin Nutr. 2004;80(1):193–8. **<u>PubMed</u> | <u>Google Scholar</u>**

6. Dat TQ, Le Nguyen THG, Loan T, Van Toan V. The prevalence of malnutrition based on anthropometry among primary schoolchildren in Binh Dinh province, Vietnam. AIMS Public Health. 2018;5(3):203–16. **<u>PubMed</u> | <u>Google Scholar</u>**

7. De Onis M, Blössner M, Borghi E, Morris R, Frongillo EA. Methodology for estimating regional and global trends of child malnutrition. Int J Epidemiol. 2004;33(6):1260–70. **<u>PubMed</u> | <u>Google Scholar</u>**

8. Kasirye I. What are the successful strategies for reducing malnutrition among young children in East Africa? Human Development Report Office (HDRO), United Nations Development Programme (UNDP); 2010. **<u>Google Scholar</u>**

9. Masibo RK. Trends and determinants of malnutrition among children age 0–59 months in Kenya (DHS 1993, 2003 and 2008–09). DHS Working Papers No. 89. ICF International; 2013. **<u>Google Scholar</u>**

10. Masiye F, Chama C, Chitah B, Jonsson D. Determinants of child nutritional status in Zambia: an analysis of a national survey. Zambia Soc Sci J. 2010;1(1):Article 4. **Google Scholar**

11. Musenge EM, Tembo S, Hankwebe M, Kahinga N, Mushibwe O, Mulenga I, et al. Prevalence and determinants of malnutrition among under-five children in Lusaka urban, Zambia. Tanzan J Health Res. 2020;21(1):1–13. 10.4314/thrb.v21i1.5 **Google Scholar**

12. Rachmi CN, Agho KE, Li M, Baur LA. Stunting, underweight and overweight in children aged 2.0–4.9 years in Indonesia: prevalence trends and associated risk factors. PLoS One. 2016;11(5):e0154756. **PubMed | Google Scholar**

13. Rahman A, Biswas SC. Nutritional status of under-5 children in Bangladesh. South Asian J Popul Health. 2009;2(1):1–11. **Google Scholar**

14. Roberfroid D, Pelto GH, Kolsteren P. Maternal comprehension of growth charts worldwide. Trop Med Int Health. 2007;12(9):1074–86. **PubMed | Google Scholar**

15. UNICEF. Tracking progress on child undernutrition. 2nd ed. Washington DC: World Bank; 2009. **Google Scholar**

16. UNICEF, WHO, World Bank. UNICEF-WHO-World Bank Joint child malnutrition estimates. New York: UNICEF; Geneva: WHO; Washington DC: World Bank; 2015. **Google Scholar**

17. UNICEF, WHO, World Bank, UN. Levels and trends in child mortality: report 2014 estimates developed by the UN Inter-agency Group for Child Mortality Estimation (UN IGME). New York: UNICEF; 2014. **Google Scholar**

18. Zambia Statistics Agency, Ministry of Health (MOH) Zambia, and ICF. Zambia Demographic and Health Survey 2018. Lusaka, Zambia and Rockville, Maryland, USA: Zambia Statistics Agency, Ministry of Health, and ICF; 2019. **Google Scholar**

